# Timing of Menarche and Menopause and Epigenetic Aging among U.S. Adults: Results from the National Health and Nutrition Examination Survey 1999-2002

**DOI:** 10.1101/2024.12.19.24319271

**Authors:** Saher Daredia, Dennis Khodasevich, Nicole Gladish, Hanyang Shen, Jamaji C. Nwanaji-Enwerem, Anne K. Bozack, Belinda L. Needham, David H. Rehkopf, Julianna Deardorff, Andres Cardenas

**Author notes:** Corresponding Author: Andres Cardenas, Department of Epidemiology and Population Health, Stanford University, Stanford, CA 94305, USA. These authors have contributed equally to this work and share senior authorship (Julianna Deardorff, Andres Cardenas).

## Abstract

Reproductive aging, including timing of menarche and menopause, influences long-term morbidity and mortality in women, yet underlying biological mechanisms remain poorly understood. Using DNA methylation-based biomarkers, we assessed associations of age at menarche (N=1,033) and menopause (N=658) with epigenetic aging in a nationally representative sample of women ≥50 years. Later age at menopause was associated with lower GrimAge epigenetic age deviation (*B* = -0.10 years, 95% CI: -0.19, -0.02). No associations were observed for menarche timing. This suggests a connection between earlier menopause and biological aging, with potential clinical implications for identifying those at high risk for age-related disease.

## INTRODUCTION

Reproductive health is not only critical to women’s overall health and wellbeing, but is increasingly shown to be a key predictor of future chronic disease risk. An earlier age of menarche, defined as first occurrence of menstruation, is associated with earlier all-cause mortality and increased risk of breast and endometrial cancers, diabetes, and cardiovascular disease (CVD).(1,2) Research on the long-term impacts of menopausal timing, defined as occurring 12 months after a woman’s last menstrual period, has varied by health outcome, with earlier onset linked to a higher risk of CVD and all-cause mortality, but a lower risk of breast cancer.(2,3)

In the past 60 years, the mean age of menarche has decreased by 0.8 years, while the mean age of natural menopause has increased by 1.5 years among women in the United States (U.S.).(3) These shifting trends in reproductive timing may have broader implications for aging processes and long-term health outcomes. Reproductive aging has also been shown to influence overall aging processes in organisms at the cellular, tissue, organ, and system levels.(4) Despite this, there is a notable gap in research exploring the relationship between reproductive timing and epigenetic age biomarkers, which are DNA methylation (DNAm)-based predictors of morbidity and mortality. Referred to as epigenetic clocks, these biomarkers are strong predictors of chronological age and derived from DNAm levels of specific cytosine-phosphate-guanine (CpG) sites.(5) Developed using different human tissue types and age-related outcomes, these clocks capture different aspects of biological aging.(5)

There are established associations between many lifestyle and environmental factors and altered epigenetic aging.(5) Only a handful of studies have considered the relationship between menarche and menopausal timing with epigenetic aging in later adulthood. Earlier menarche has been linked to accelerated epigenetic aging in studies of adolescent and premenopausal women.(6,7) Additionally, later menopause has been associated with slower epigenetic aging in a few reports.(8,9) However, most of these studies are limited by small sample sizes and limited diversity in participant characteristics, reducing generalizability of findings to the broader U.S. population.

In this study, we examine the association of ages at menarche and menopause with epigenetic aging in the National Health and Nutrition Examination Survey (NHANES), a nationally representative sample of adults in the United States that includes demographic, questionnaire, and laboratory data. We hypothesize that earlier timing of these reproductive phases is associated with accelerated epigenetic aging.

## METHODS

NHANES is a biannual, nationally representative cross-sectional survey conducted by the National Center for Health Statistics (NCHS) to assess the health of the noninstitutionalized U.S. population.(10) Our study consisted of adult participants ages 50-84 years who participated in the 1999-2000 and 2001-2002 NHANES cycles and provided blood samples for DNAm analysis. After applying eligibility criteria, we analyzed N=1,033 and N=658 participants who had self-reported questionnaire data available on timing of menarche and menopause, respectively.

Further details on defining the analytic sample can be found in **Supplemental Figures 1-2**. Ages at menarche and menopause were self-reported in the NHANES reproductive health questionnaire which was administered to female-identifying participants.(10) Data on epigenetic age and DNAm-derived blood cell proportion estimates were downloaded from the NHANES website.(10) Our analysis included several epigenetic age measures, including the Horvath, Hannum, SkinBlood, PhenoAge, GrimAge, GrimAge2, DunedinPoAm, and DNAmTL clocks.(5) Epigenetic age deviation was calculated for each clock as the residuals from a linear regression model of chronological age on each epigenetic aging biomarker. Positive epigenetic age deviation (>0 years) occurs when epigenetic age is greater than chronological age, reflecting accelerated epigenetic aging; meanwhile, negative epigenetic age deviation (<0 years) indicates epigenetic age is lower than chronological age. DNAmTL, an estimate of leukocyte telomere length, and DunedinPoAm, an estimate of relative pace of aging, were left untransformed in our analysis. Generalized linear regression models with survey weights were used to evaluate associations of ages at menarche and menopause with each epigenetic age deviation measure.

Main models were adjusted for chronological age in years (continuous), chronological age in years squared (continuous), and self-identified race/ethnicity. Further details on exposure and outcome assessment, covariates, and sensitivity analyses can be found in the **Supplemental Methods**.

## RESULTS

Table 1 describes the unweighted characteristics of study participants in the menarche and menopause analytic samples. The majority of participants identified as Non-Hispanic White (40% of both samples), followed by Mexican American (29% of both samples), Non-Hispanic Black (menarche sample: 21%; menopause sample: 19%), Other Hispanic (menarche sample: 6%; menopause sample: 7%), and Other or Multiracial (4% of both samples). Participants in the menarche and menopause samples had a mean (SD) chronological age of 64.9 (9.3) and 66.1 (9.1) years, respectively, at the time of the NHANES screening. In the menarche sample, participants reported reaching menarche at an mean age of 12.9 (1.7) years. In the menopause sample, participants reported experiencing menopause at an mean age of 50.0 (4.9) years.

**Table 1:** Unweighted participant characteristics included in analyses of age at menarche and menopause: NHANES 1999-2002. Values represent unweighted count (%) or mean (SD).

|  | **Menarche Sample**<br>**(N=1,033)** | **Menopause Sample**<br>**(N=658)** |
| --- | --- | --- |
| **Age at screening, years** | 64.9 (9.3) | 66.1 (9.1) |
| **Age at menarche, years** | 12.9 (1.7) | 13.0 (1.7) |
| **Age at menopause, years** | 49.9 (4.9)* | 50.0 (4.9) |
| **Race/ethnicity** |  |  |
| Mexican American | 300 (29.0%) | 193 (29.3%) |
| Other Hispanic | 66 (6.4%) | 49 (7.4%) |
| Non-Hispanic White | 410 (39.7%) | 264 (40.1%) |
| Non-Hispanic Black | 216 (20.9%) | 124 (18.8%) |
| Other Race/Multi-Racial | 41 (4.0%) | 28 (4.3%) |
| **Body mass index (BMI) at screening, kg/m<sup>2</sup>** | 29.4 (6.5) | 29.4 (6.3) |
| Missing (n) | 23 (2.2%) | 15 (2.3%) |
| **Smoking status at screening** |  |  |
| Ever | 418 (40.5%) | 265 (40.3%) |
| Never | 612 (59.2%) | 391 (59.4%) |
| Missing | 3 (0.3%) | 2 (0.3%) |
| **Alcohol intake at screening** |  |  |
| Abstainer | 557 (53.9%) | 350 (53.2%) |
| Moderate drinker | 450 (43.6%) | 293 (44.5%) |
| Heavy drinker | 25 (2.4%) | 14 (2.1%) |
| Missing | 1 (0.1%) | 1 (0.2%) |

\*Available for N=646 of N=1,033 who met the criteria for having undergone menopause.

Participants in both samples had a mean body mass index (BMI) of 29.4 (menarche SD: 6.5; menopause SD: 6.3) kg/m^2^ at the time of screening. Most participants reported never smoking (59% of both samples) and abstaining from alcohol (menarche sample: 54%; menopause sample: 53%).

We observed strong positive correlations between chronological age and epigenetic age across all unique 1,045 participants in both samples, ranging from *r*=0.76 (Median Absolute Error, MAE: 10.7 years) with PhenoAge to *r*=0.89 (MAE: 2.8 years) with SkinBlood (**Supplemental Figure 3**). DNAmTL was negatively correlated with chronological age (*r*=- 0.60), indicating shortening telomere length with increasing age. The DunedinPoAm biomarker was uncorrelated with chronological age, as expected.

In primary models adjusted for chronological age, chronological age squared, and self-identified race/ethnicity, we did not observe any associations between age at menarche and epigenetic age deviation across any of the epigenetic aging biomarkers (**Figure 1**). Additionally, there were no observed associations between ages at menarche and menopause with untransformed DNAmTL and DunedinPoAm (**Supplementary Figure 4**). However, one-year greater age at menopause was significantly associated with lower GrimAge epigenetic age deviation (*B* = -0.10 years, 95% CI: -0.19, -0.02) at the time of the NHANES screening. A similar trend was observed for GrimAge2, where increased age at menopause was associated with a negative deviation (*B* = -0.10 years, 95% CI: -0.20, 0.00), but this association was imprecise. Given these findings, we examined associations of age at menopause with the DNAm predicted components of GrimAge and GrimAge2. We observed that one-year greater age at menopause was associated with lower DNA methylation estimates of adrenomedullin (ADM) (*B* = -0.36, 95% CI: -0.68, -0.04) and plasminogen activation inhibitor 1 (PAI1) (*B* = -56.70, 95% CI: -113.22, -0.19) — both of which are components of the GrimAge and GrimAge2 biomarkers.

**Figure 1:**
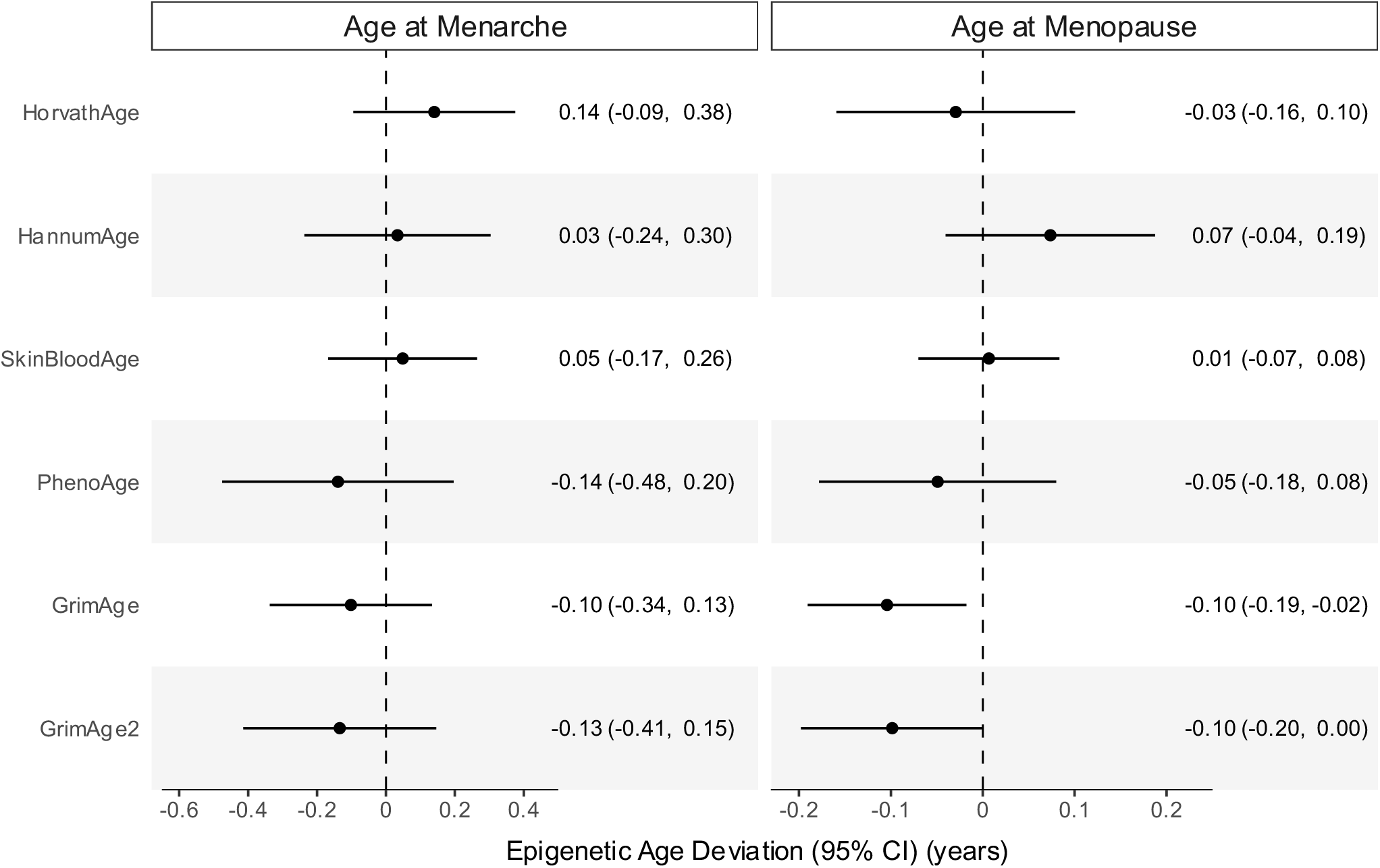
Adjusted estimates and 95% CI for associations of epigenetic age deviation with (a) age at menarche and (b) age at menopause.

Associations between age at menopause and epigenetic age deviation from the other clocks were consistently close to the null (**Figure 1**). Sensitivity analyses additionally adjusted for (i) BMI, smoking status, and alcohol intake at the time of the NHANES screening and (ii) estimated cell-type proportions showed similar, but attenuated, associations between later age at menopause and negative GrimAge and GrimAge2 epigenetic age deviation (**Supplemental Figures 5-6**).

## DISCUSSION

We examined associations of menarche and menopausal timing with epigenetic aging biomarkers in a representative sample of United States women aged 50-84 years from NHANES. In models adjusted for chronological age and race/ethnicity, we observed that greater age at menopause was associated with lower GrimAge. We did not find any relationship between age at menarche and epigenetic aging.

Our findings on menopausal timing are supported by existing literature. Another study also found an inverse relationship between age at menopause and GrimAge epigenetic aging (*B*=-0.06 years) among women greater than 50 years of age.(9) GrimAge is a composite biomarker that is based on DNAm surrogates of seven plasma proteins linked to morbidity and mortality as well as smoking pack years (i.e., the number of packs of cigarettes smoked per day multiplied by the numbers of years an individual smoked).(9) Unlike first-generation epigenetic clocks, which were trained to predict chronological age, GrimAge is a robust predictor of time-to-disease and time-to-death.(9) Since GrimAge is associated with disease risk and mortality, the observed association likely reflects the long-term health impacts of menopausal timing which influences biological aging processes and susceptibility to age-related chronic disease. This is supported by our similar findings with GrimAge2, an updated version that leverages additional DNAm-based estimators of plasma proteins and better predicts mortality across multiple racial/ethnic groups. A separate meta-analysis did report similar findings with menopausal timing and Horvath epigenetic aging across three predominantly White U.S and European-based cohorts (*B*=-0.01 to -0.06 years) (8). While the overall estimate was significant, associations were not significant among individual cohorts. Nonetheless, the results were in the same direction as those in NHANES, and the findings with the Horvath epigenetic aging biomarker suggest that other biological aging pathways may be involved in menopausal timing, although this was not observed in our sample.

In sensitivity analyses, we found that our results on menopausal timing and epigenetic aging were attenuated after adjustment for BMI, smoking, and alcohol intake. While these covariates are potential confounders of the association between menopausal timing and epigenetic aging, they were ascertained at the time of the NHANES screening/sample collection (post exposure) and no retrospective data was available. Since assessment occurred after menopause, it is possible that these covariates are on the causal pathway in the relationship, in which case adjustment may have led to overadjustment bias. We also observed attenuation after adjusting for estimated cell-type proportions, suggesting that the link between menopausal timing and GrimAge may be influenced by inflammatory or other leukocyte-mediated processes that occur as the immune system ages.

Notably, our null findings regarding age at menarche differ from those reported in previous studies. A study of primarily White women based in the San Francisco Bay Area found that later age at menarche was associated with negative GrimAge epigenetic age deviation (*B*=- 0.45 years) among premenopausal women ages 25 to 51 years.(6) Another longitudinal study observed that a five-year positive Horvath epigenetic age deviation was associated with significantly earlier age of menarche (Hazard Ratio, HR=1.29) among Chilean adolescent girls.

A possible explanation for the discrepancy between these findings and ours is the more proximal timing between menarche and epigenetic age assessment in these studies. In contrast, the longer time gap between menarche and assessment of epigenetic age in our sample of women aged >=50 years, coupled with the influence of other exposures that may have altered epigenetic aging trajectories, may have contributed to the lack of a detectable association.

To our knowledge, this is the largest study of menarche and menopausal timing and epigenetic aging in a sample of adults that is representative of the entire U.S. population. Still, our study does have some important limitations. Although we adjusted for demographic and behavioral covariates, there may be residual confounding due to unmeasured covariates such as pre-pubertal and pre-menopausal physical activity, diet, BMI, smoking, and alcohol consumption. Our exposures, ages at menarche and menopause, may also be subject to recall error, but we expect this to be non-differential and bias our results towards the null. Future research should examine longitudinal changes in epigenetic aging before and after the pubertal and menopausal transitions. Additionally, more objective clinical indicators, such as Tanner staging, can be used to better assess the relationship between pubertal timing and epigenetic aging as menarche often occurs later in the pubertal transition. Regardless, our findings do suggest that earlier timing of reproductive phases, particularly menopause, is associated with accelerated epigenetic aging, which may have clinical implications for early identification of those at high risk for age-related diseases.

## Data Availability

All data produced are available online at https://wwwn.cdc.gov/nchs/nhanes/default.aspx.

## DECLARATIONS

### Ethics approval and consent to participate

All NHANES participants provided written informed consent and study protocols were approved by the NCHS Research Ethics Review Board.

### Consent for publication

Not applicable.

### Availability of data and materials

All data used for this analysis is publicly available from the NHANES website (https://www.cdc.gov/nchs/nhanes/index.htm).

### Authors’ contributions

SD conceived of the analyses, performed data analysis, visualization, and original writing. DK, NG, HS, JCN, AKB, BLN, and DHR contributed to the analysis and edited the manuscript. JD and AC conceived of the analyses, supervised the work, and contributed to writing/editing of the manuscript. All authors reviewed and approved the final manuscript.

## Acknowledgements

We gratefully acknowledge the NHANES study staff and participants for their contributions.

## Appendix

Associations were assessed using survey-weighted generalized linear regression models. All models were adjusted for chronological age in years (continuous), chronological age squared, and self-identified race/ethnicity. Estimates are regression coefficients (*B*) that represent the estimated change in each epigenetic age biomarker per one-year increase in age at menarche or age at menopause.

## Supplemental Methods

### Age at Menarche and Menopause

Age at menarche was obtained by asking participants, "How old were you when you had your first menstrual period?" (Reproductive Health Questionnaire, RHQ010).(1,2)

Age at menopause was assessed through a series of questions in the RHQ.(1,2) Participants were first asked “Have you had regular periods in the last 12 months?” (RHQ030). Those answering “No” due to medical conditions/treatment or usual irregularity (N=109), “Yes” (N=17), or unknown period regularity (N=1) were excluded (RHQ040).

Participants who had their last menstrual period <12 months since the time of screening (N=43) (RHQ050) and those who did not know the time elapsed since their last menstrual period (N=1, RHQ050) or age at last period (N=75, RHQ060) were further excluded. Finally, those reporting menopause before age 40 (N=174) or after age 62 years (N=4) were removed since pathologic conditions may have influenced the occurrence of menopause at such ages.(3) Age at menopause was then calculated as 12 months after age at last menstrual period for those remaining in the sample.

### DNA Methylation and Epigenetic Age

Epigenetic age measures and DNA methylation (DNAm)-based leukocyte proportion estimates were downloaded from the NHANES website.(4) DNA was extracted from whole blood of a subset of NHANES participants aged ≥50 years old from the 1999-2000 and 2000-2001 survey cycles. Genome-wide DNAm was measured using the Illumina EPIC BeadChip array. Quality control steps and DNAm data processing were completed, as described elsewhere.(4) The Horvath,(5) Hannum,(6) SkinBlood,(7) PhenoAge,(8) GrimAge,(9) GrimAge2,(10) DNAmTL,(11) and DunedinPoAm(12) epigenetic age biomarkers were included in analyses. Pearson correlation coefficients and median absolute error (MAE) were used to assess the fit for each epigenetic age biomarker and chronological age.

### Statistical Analysis

Generalized linear regression models were run using the *svyglm* function from the R ‘Survey’ package(13) to account for survey weights, correcting for selection, non-response, and coverage biases, following NHANES analytic guidelines for the epigenetic clock sample. Potential confounders/model covariates were identified *a priori* and included chronological age in years (continuous), chronological age in years squared (continuous), and self-identified race/ethnicity (Non-Hispanic White, Mexican American, Other Hispanic, Non-Hispanic Black, Other Race - Including Multi-Racial). We performed sensitivity analyses further adjusting models for the original covariate set plus (i) body mass index (BMI) (continuous), smoking status (ever, never), and alcohol intake (abstainer, moderate, heavy drinker) at the time of the NHANES screening and (ii) estimated cell-type proportions (CD8 cells, CD4 cells, NK cells, B cells, monocytes, and neutrophils). Finally, given the main epigenetic age results, we evaluated associations of age at menopause with the DNA methylation components of GrimAge and GrimAge2 — smoking pack-years, adrenomedullin (ADM), beta-2 microglobulin (B2M), cystatin C, growth differentiation factor 15 (GDF15), leptin, plasminogen activation inhibitor 1 (PAI1), tissue inhibitor metalloproteinase 1 (TIMP1), C-reactive protein (CRP), and hemoglobin A1C.(9,10) We report adjusted regression coefficients (*B*) and 95% Confidence Intervals (CIs) to evaluate statistical significance and precision. All statistical analyses were performed using R version 4.3.1.(14)

---

**Supplemental Figure 1:**
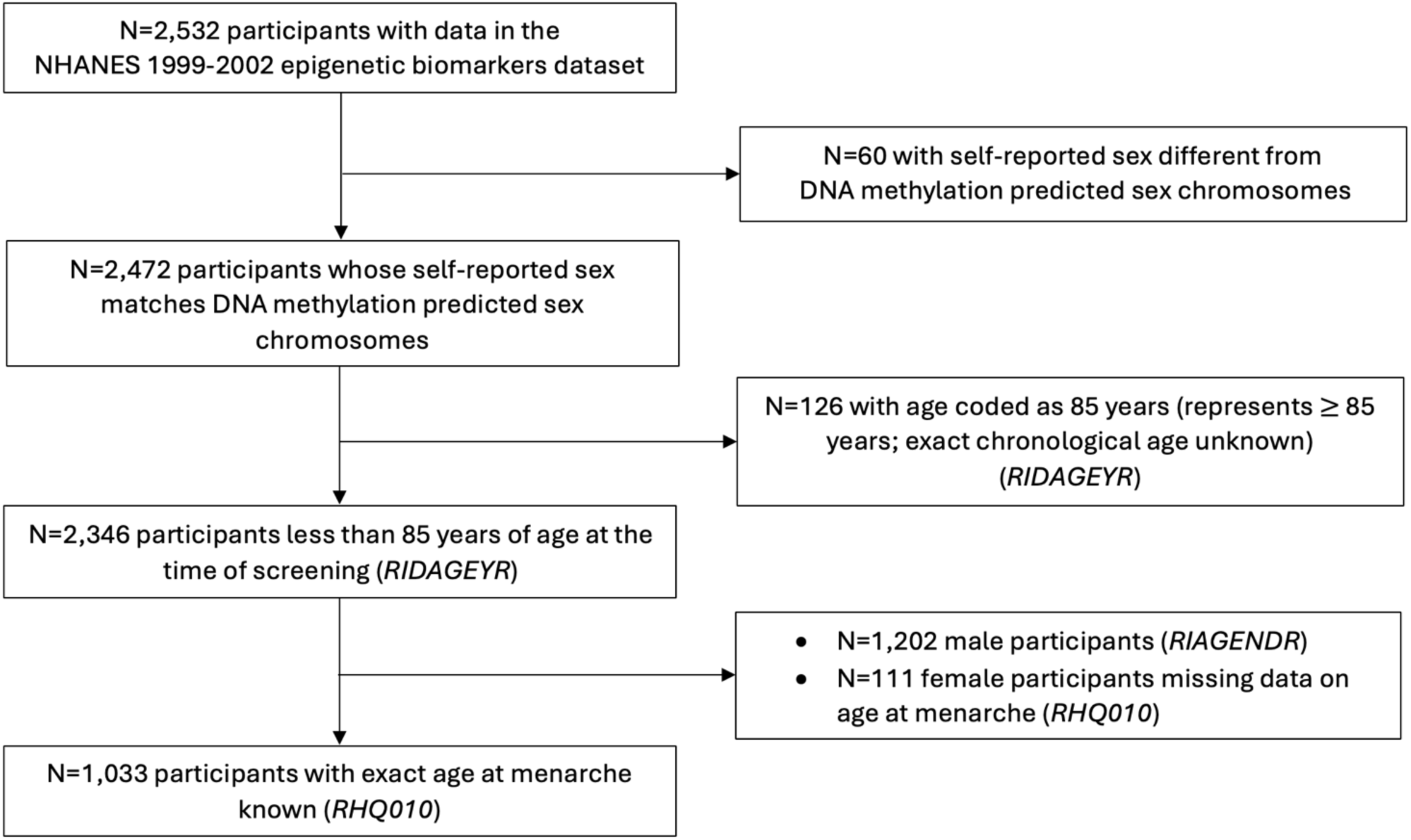
Flowchart of menarche analysis participants (N=1,033).

**Supplemental Figure 2:**
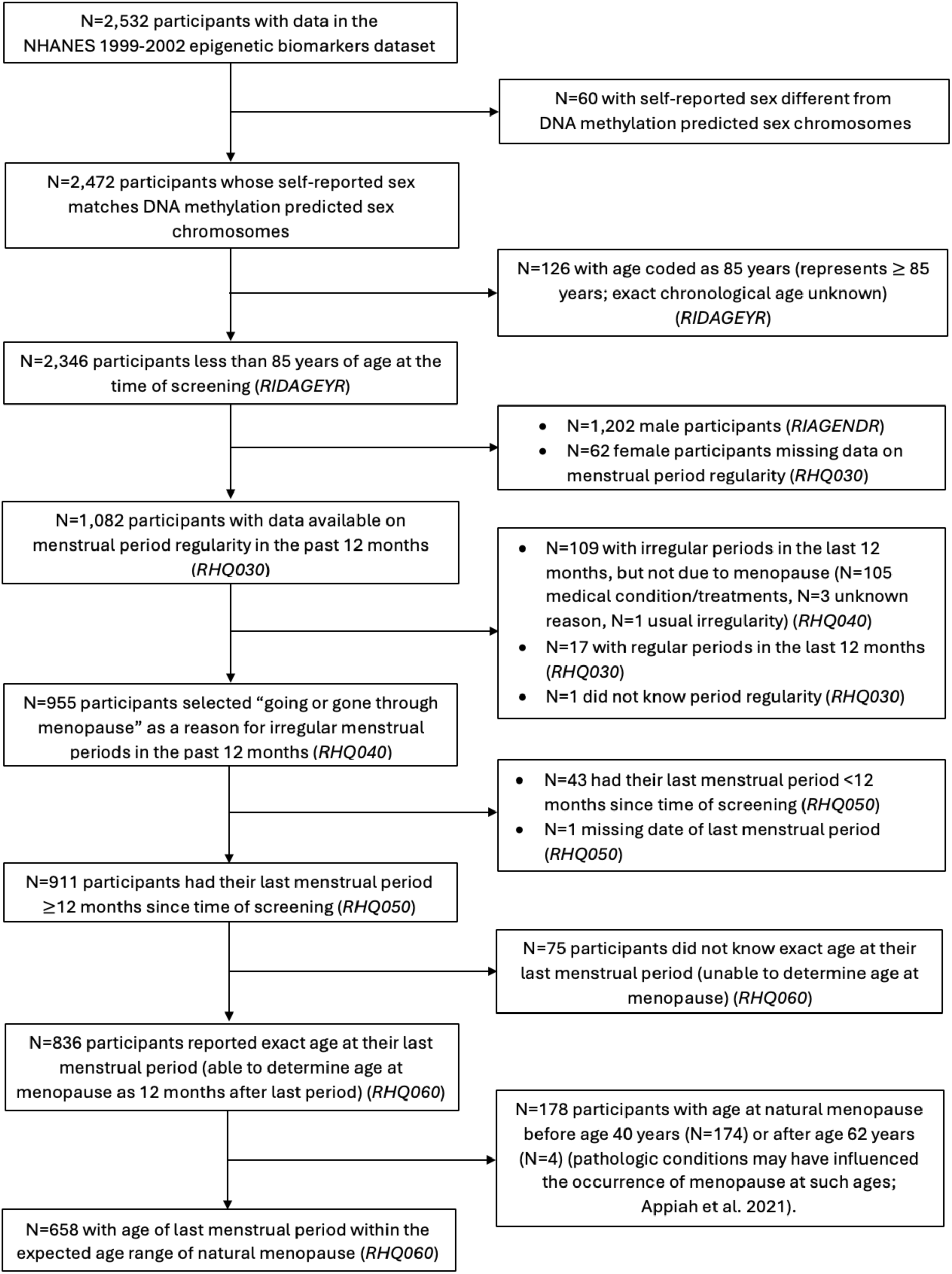
Flowchart of menopause analysis participants (N=658).

**Supplemental Figure 3:**
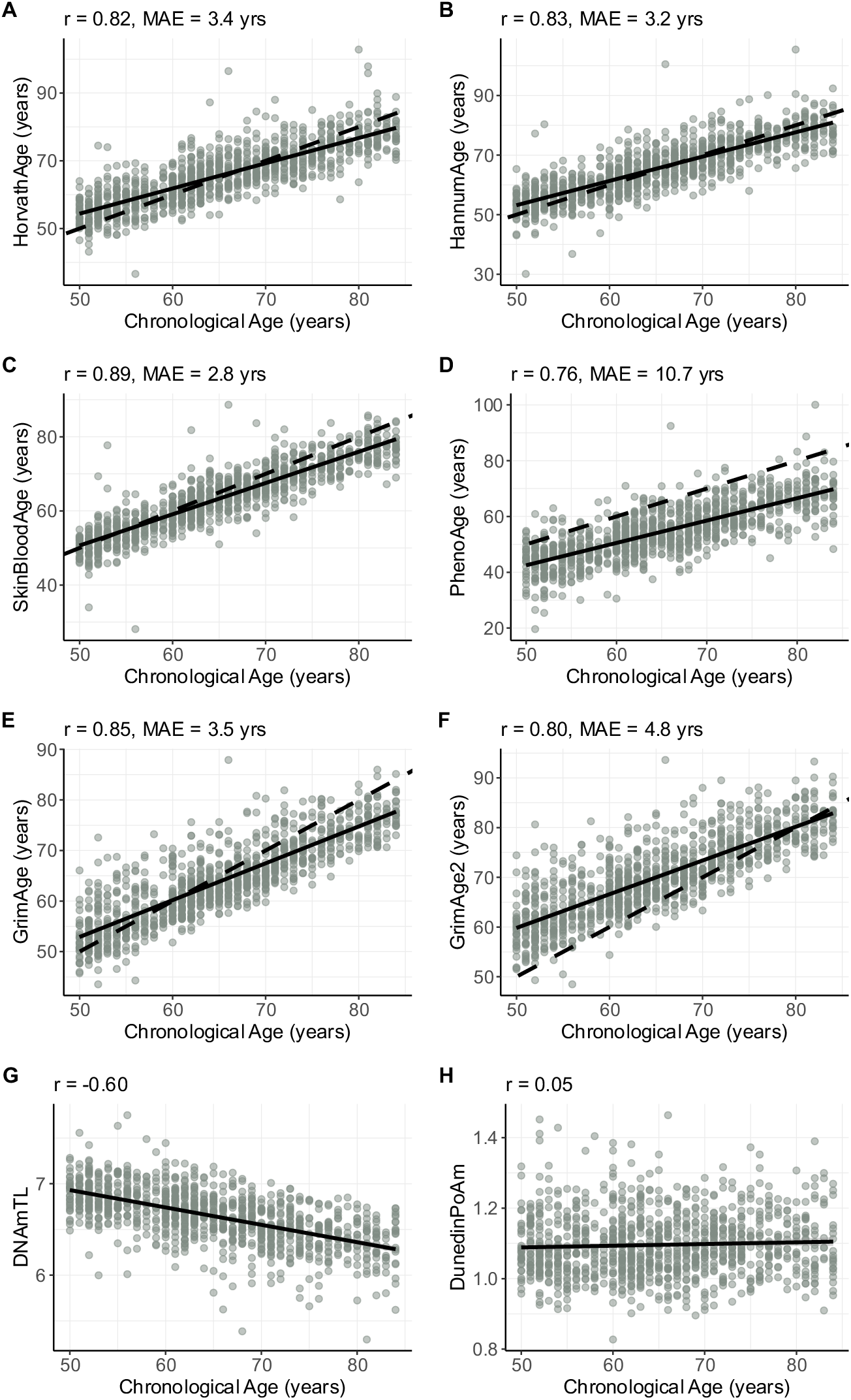
Epigenetic clock performance in total sample (both menarche and menopause analyses) (N=1,045).

Pearson correlation coefficient *r* and median absolute error (MAE) between chronological age based on birth date and epigenetic age estimated by each epigenetic clock.

The linear trendline and 95% CI are plotted as a solid line with shaded area and the identity line (y=x) is plotted as a dashed line.

**Supplemental Figure 4:**
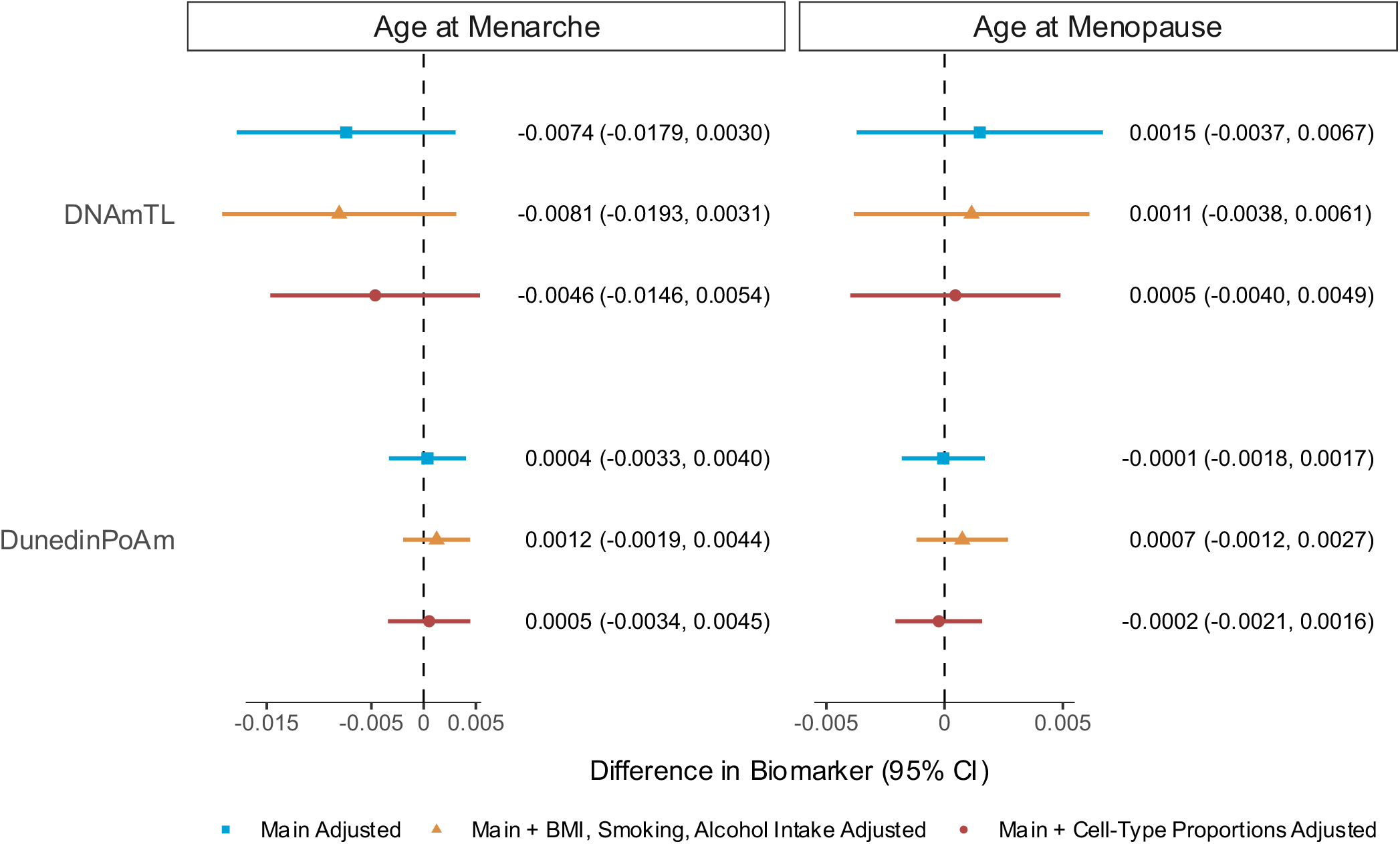
Associations of DNAmTL and DunedinPoAm with (a) age at menarche and (b) age at menopause.

Associations were assessed using survey-weighted generalized linear regression models. Main models (blue) were adjusted for chronological age in years (continuous), chronological age squared, and self-identified race/ethnicity. In sensitivity analyses, models were adjusted for covariates in main models in addition to i) BMI, smoking status, and alcohol intake at NHANES screening (yellow) and ii) cell-type proportions (red).

Estimates are regression coefficients (*B*) that represent the estimated change in each biomarker per one-year increase in age at menarche or age at menopause.

**Supplemental Figure 5:**
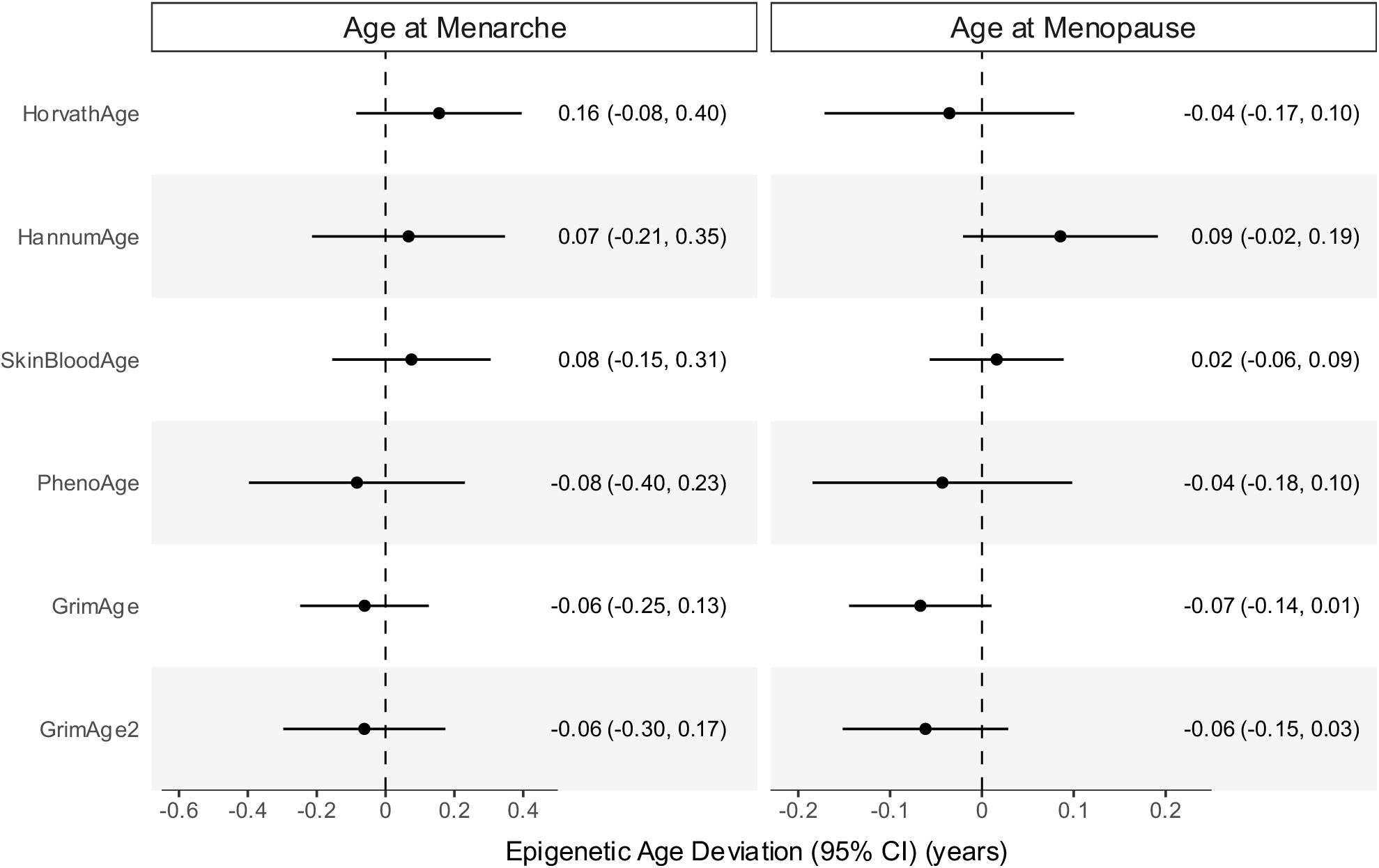
Associations of epigenetic age deviation with (a) age at menarche and (b) age at menopause (additionally adjusted for BMI, smoking, and alcohol intake).

Associations were assessed using survey-weighted generalized linear regression models. All models were adjusted for chronological age in years (continuous), chronological age squared, self-identified race/ethnicity, body mass index at time of screening, smoking status at time of screening, and alcohol intake at time of screening.

Estimates are regression coefficients (*B*) that represent the estimated change in each epigenetic age biomarker per one-year increase in age at menarche or age at menopause.

**Supplemental Figure 6:**
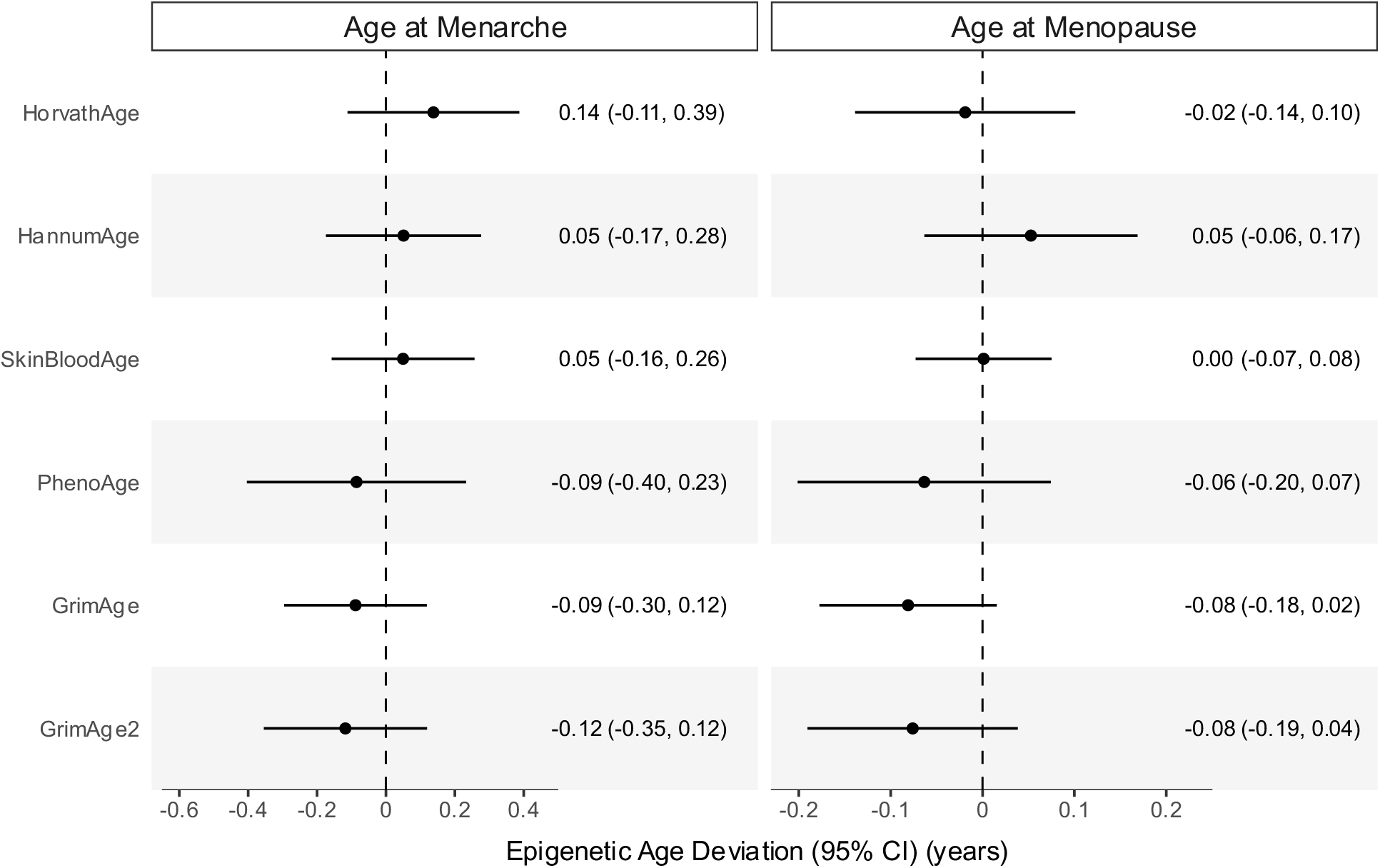
Associations of epigenetic age deviation with (a) age at menarche and (b) age at menopause (additionally adjusted for cell-type proportions).

Associations were assessed using survey-weighted generalized linear regression models. All models were adjusted for chronological age in years (continuous), chronological age squared, self-identified race/ethnicity, and estimated cell-type proportions (CD8 cells, CD4 cells, NK cells, B cells, monocytes, and neutrophils).

Estimates are regression coefficients (*B*) that represent the estimated change in each epigenetic age biomarker per one-year increase in age at menarche or age at menopause.

